# Methodological Considerations in Sibling Analyses of Prenatal Acetaminophen

**DOI:** 10.64898/2026.03.27.26349515

**Authors:** Viktor H. Ahlqvist, Hugo Sjöqvist, Renee M. Gardner, Brian K. Lee

**Author notes:** Corresponding author: Brian Lee.; 3215 Market St, Philadelphia, PA 19104.

## Abstract

**Background:** Sibling-matched designs control for shared familial confounding but remain vulnerable to non-shared confounders. Bi-directional sensitivity analyses, which stratify families by whether the older or younger sibling was exposed, are commonly used to assess carryover effects. We aimed to demonstrate how this methodological approach can introduce severe confounding by parity.

**Methods:** We conducted simulations motivated by a recent epidemiological study. The true causal effect of a hypothetical exposure (prenatal acetaminophen) on neurodevelopmental outcomes was set to strictly null. To introduce parity-related confounding, baseline exposure and outcome probabilities were varied slightly by birth order. We compared conditional logistic regression effect estimates from total sibling models against bi-directional stratified models.

**Results:** In the total simulated sibling cohort, models yielded the true null effect (odds ratio = 1.00) when adjusting for parity. However, the bi-directional analyses exhibited divergent artifactual signals. Because parity is perfectly collinear with exposure in these stratified subsets, it cannot be adjusted for. For example, when the older sibling was exposed, the odds ratio for autism spectrum disorder was 1.68; when the younger was exposed, the odds ratio was 0.60.

**Conclusions:** Divergent estimates in bi-directional sibling analyses can be a predictable artifact of parity confounding rather than evidence of carryover effects or invalidating unmeasured bias. Overall sibling models adjusting for parity may remain robust despite divergent stratified sensitivity results.

## Introduction

Sibling analyses effectively address familial confounding (e.g. from genetics or parental health) in observational studies.^1^ In a Taiwanese sample, Lee et al. observed that positive associations between prenatal acetaminophen and neurodevelopmental outcomes became null in sibling analyses.^2^ These null results are consistent with findings from Norway, Sweden, and Japan.^3-5^ However, citing a sensitivity analysis where risks flipped depending on whether the older or younger sibling was exposed (e.g., ADHD hazard ratios of 1.33 vs. 0.75 respectively), Lee et al. concluded that unaddressed biases could invalidate their sibling design. Here, we show this divergence is a predictable artifact of stratifying on a confounder perfectly collinear with exposure, rather than an unaddressed bias invalidating their null finding.

## Methods

The validity of sibling analyses relies on the assumption that the exposure and outcome status of an earlier pregnancy do not influence the subsequent pregnancy (termed “carryover effects”).^6^ To assess the presence of carryover effects, a sensitivity analysis (“bi-directional analysis”) compares results between families stratified by which sibling was exposed. This entails separate models for pairs where exposure occurs in the first-versus second-born child, and vice-versa, aligning exposure with birth order. Consistent results suggest that bias from carryover effects is absent. However, divergent results do not necessarily imply bias in the main sibling analysis; it could reflect bias inherent to the sensitivity analysis itself. Specifically, bi-directional analysis is highly sensitive to confounding by parity (i.e., birth order effects). Because bi-directional subsets are strictly discordant for both exposure and birth order, parity becomes perfectly collinear with exposure, making adjustment impossible. Notably, parity is appropriately adjusted for in Lee et al.’s main sibling analysis but necessarily omitted from bi-directional models.

To demonstrate how parity-related confounding artificially generates divergence in bi-directional analyses, we simulated a hypothetical sibling cohort. The true causal effect of prenatal acetaminophen exposure on neurodevelopment was null, but exposure and outcome probabilities varied slightly by birth order.

We generated 500 cohorts of 525,000 two-sibling families. A normally distributed latent factor simulated shared familial confounding, influencing both acetaminophen exposure and ASD/ADHD probabilities. Exposure and outcome status were generated using binomial distributions. To introduce parity confounding, we applied a slight drift between siblings. Consistent with Lee et al.’s observation of no substantial change in exposure prevalence during the study, exposure probability was 48% for the first sibling and 50% for the second sibling. Using parameters reflecting birth-order effects for neurodevelopmental diagnoses in Taiwan,^7^ ASD probability was 1.25% for the first sibling and 0.75% for the second. ADHD probability was 6% for the first sibling and 4.5% for the second. The true causal effect was null for all individuals.

We restricted analyses to exposure-discordant families. Using conditional logistic regression grouped by family, we examined four model specifications:

1. All families, unadjusted for parity.
2. All families, adjusting for parity.
3. Bi-directional analysis: first child exposed.
4. Bi-directional analysis: second child exposed.

Odds ratios (OR) with empirical 95% CI were calculated. See Supplement for Stata MP 19.5 code.

## Results

In the total sibling cohort, omitting parity adjustment introduced minimal bias (e.g. ASD OR=0.98 unadjusted vs. OR=1.00 adjusted) (**Table)**. However, bi-directional analyses, where parity perfectly aligns with exposure and therefore cannot be adjusted for, exhibited divergent signals. When the older sibling was exposed, ASD OR=1.68. When the younger sibling was exposed, ASD OR=0.60. These results mirror the magnitude of Lee et al.’s estimates (hazard ratios=1.75 and 0.74, respectively). Similar artifactual divergence occurred for ADHD.

## Discussion

Divergent bi-directional sibling analysis results, like those observed by Lee et al., can arise from confounders not shared between siblings. Because parity perfectly aligns with exposure in bi-directional analysis, it strongly confounds this sensitivity analysis while minimally affecting the main sibling analysis.

Adjusting for birth year in bi-directional analysis does not resolve parity confounding as they are distinct, non-shared confounders.^8,9^ Birth year adjustment controls for secular trends (e.g., shifting diagnostic criteria). In contrast, parity adjustment accounts for biological and intra-familial changes (e.g., maternal physiological differences across pregnancies). Because a firstborn child can be born in any calendar year, adjusting solely for birth year leaves the bi-directional model vulnerable to confounding by parity.

Our simulation shows even minor parity effects on exposure and outcome probabilities can generate substantial bias in bi-directional analysis. While consistent bi-directional analysis results support the validity of sibling analyses, divergent bi-directional analysis results do not necessarily invalidate sibling analyses. The divergence may simply reflect parity confounding inherent to the sensitivity analysis, and not a bias in the main sibling analysis itself. Consequently, Lee et al.’s overall null sibling findings, which appropriately adjusted for parity, are likely more robust than their bi-directional analyses suggest.

**Table:** Associations of prenatal acetaminophen exposure and ASD and ADHD in simulation analyses in comparison to results from Lee et al.

|  | True effect of exposure on outcome | Total sibling sample (not adjusted for parity) | Total sibling sample (adjusted for parity) | Bi-directional sibling analysis: older sibling exposed, younger sibling unexposed | Bi-directional sibling analysis: older sibling unexposed, younger sibling exposed | Mean number of retained pairs in total sibling sample |
| --- | --- | --- | --- | --- | --- | --- |
| **Simulation odds ratios and 95% CI** |  |  |  |  |  |  |
| ASD | 1.00 | 0.98<br>(0.93, 1.03) | 1.00<br>(0.95, 1.05) | 1.68<br>(1.56, 1.81) | 0.60<br>(0.56, 0.64) | 192,372 |
| ADHD | 1.00 | 0.99<br>(0.96, 1.01) | 1.00<br>(0.98, 1.03) | 1.36<br>(1.30, 1.41) | 0.74<br>(0.71, 0.76) | 192,372 |
| **Lee et al. hazard ratios and 95% CI** |  |  |  |  |  |  |
| ASD | N/A | Not reported | 0.98<br>(0.90, 1.07) | 1.75<br>(1.29, 2.36) | 0.74<br>(0.57, 0.96) | 193,609 |
| ADHD | N/A | Not reported | 0.99<br>(0.96, 1.03) | 1.33<br>(1.17, 1.52) | 0.75<br>(0.67, 0.84) | 192,712 |

## Supporting information

Stata code

## Data Availability

All data produced in the present work are contained in the Supplement.

## Data availability statement

All data produced in the present work are contained in the Supplement.

## References

1. Ahlqvist VH, Lee BK, Chiu Y-H. Sibling comparisons to account for confounding in observational studies. JAMA. 2026;

2. Lee P-C, Chuang Y-H, Hu Y-H, et al. Maternal Acetaminophen Use and Child Neurodevelopment. JAMA pediatrics. 2026;

3. Gustavson K, Ystrom E, Ask H, et al. Acetaminophen use during pregnancy and offspring attention deficit hyperactivity disorder–a longitudinal sibling control study. JCPP advances. 2021;1(2):e12020.

4. Ahlqvist VH, Sjöqvist H, Dalman C, et al. Acetaminophen use during pregnancy and children’s risk of autism, ADHD, and intellectual disability. Jama. 2024;331(14):1205–1214.

5. Okubo Y, Hayakawa I, Sugitate R, Nariai H. Maternal acetaminophen use and offspring’s neurodevelopmental outcome: a nationwide birth cohort study. Paediatric and perinatal epidemiology. 2026;40(1):70–79.

6. Sjölander A, Frisell T, Kuja-Halkola R, Öberg S, Zetterqvist J. Carryover ePects in sibling comparison designs. Epidemiology. 2016;27(6):852–858.

7. Su Y-Y, Chen C-J, Chen M-H, et al. Sibling Influence on Childhood Neurodevelopmental Disorders: A Cohort Study in Taiwan. Researchsquarecom. 2025;Preprint

8. Frisell T, Öberg S, Kuja-Halkola R, Sjölander A. Sibling comparison designs: bias from non-shared confounders and measurement error. Epidemiology. 2012;23(5):713–720.

9. Sudan M, Kheifets LI, Arah OA, Divan HA, Olsen J. Complexities of sibling analysis when exposures and outcomes change with time and birth order. Journal of Exposure Science & Environmental Epidemiology. 2014;24(5):482–488.

