## Supplementary material for "Methodological Considerations in Sibling Analyses of Prenatal Acetaminophen": Stata code

```
// Simulation to examine impact of confounding by parity on bi-directional analyses
// Ahlqvist et al., 2026
// Outcome of ADHD

clear all
set seed 123

// 1. Set up a temporary postfile to store the results of each iteration
tempfile sim_results
postfile mypost or_first or_second or_all or_adjustrank n_discordant using `sim_results', replace

display "Starting 500 iterations"

// 2. Start the timer
timer clear 1
timer on 1

// 3. Start the simulation loop
forvalues i = 1/500 {

    quietly {
        clear
        set obs 525000

        gen famid = _n
        gen u = rnormal()

        // Use expand 2 instead of reshape long
        expand 2
        bysort famid: gen sib_rank = _n

        // Generate exposure and outcome directly
        gen exposure = rbinomial(1, invlogit(logit(.48) + log(4)*u)) if sib_rank==1
        replace exposure = rbinomial(1, invlogit(logit(.50) + log(4)*u)) if sib_rank==2

        gen outcome = rbinomial(1, invlogit(logit(.06) + log(1)*exposure + log(4)*u)) if sib_rank==1
        replace outcome = rbinomial(1, invlogit(logit(.045) + log(1)*exposure + log(4)*u)) if sib_rank==2

        // Keep only discordant exposure pairs
        bysort famid: egen exp_sum = sum(exposure)
        keep if exp_sum == 1
```

```

        // Count retained pairs
        local n_pairs = _N / 2

        // Define sets for the conditional models
        bysort famid (sib_rank): gen is_set_A = (exposure[1] == 1)

        // Estimate models and store the exponentiated odds ratios
        clogit outcome exposure if is_set_A==1, or group(famid)
        local or_1 = exp(_b[exposure])

        clogit outcome exposure if is_set_A==0, or group(famid)
        local or_2 = exp(_b[exposure])

        clogit outcome exposure , or group(famid)
        local or_3 = exp(_b[exposure])

        clogit outcome exposure sib_rank, or group(famid)
        local or_4 = exp(_b[exposure])

        // Post the extracted values to the file
        post mypost (`or_1') (`or_2') (`or_3') (`or_4') (`n_pairs')
    }

    // Print progress to the console
    if mod(`i', 50) == 0 {
        noisily display "Completed iteration `i'"
    }
}

// 4. Stop the timer
timer off 1

// 5. Close the postfile
postclose mypost

// 6. Load the aggregated results and display the final summaries
use `sim_results', clear

display ""
display "--- Mean Number of Discordant Exposure Pairs Retained ---"
summarize n_discordant, meanonly
display "Mean retained pairs: " round(r(mean), 1)
display ""

display "--- Median Odds Ratios and Empirical 95% CIs (500 runs) ---"
display "---------------------------------------------------------"
display "Variable       |      2.5% |       50% |     97.5%"
```

```
display "---------------------------------------------------------"
foreach var in or_first or_second or_all or_adjustrank {
    quietly centile `var', centile(2.5 50 97.5)
    display %-14s "`var'" " | " %9.6f r(c_1) " | " %9.6f r(c_2) " | " %9.6f r(c_3)
}
display "---------------------------------------------------------"
display ""

// 7. Display the total execution time
quietly timer list 1
local minutes = floor(r(t1) / 60)
local seconds = round(mod(r(t1), 60), 0.1)
display "--- Simulation Execution Time ---"
display "Total time: `minutes' minutes and `seconds' seconds"
```

```
// Simulation to examine impact of confounding by parity on bi-directional analyses
// Ahlqvist et al., 2026
// Outcome of ASD

clear all
set seed 123

// 1. Set up a temporary postfile to store the results of each iteration
tempfile sim_results
postfile mypost or_first or_second or_all or_adjustrank n_discordant using `sim_results', replace

display "Starting 500 iterations"

// 2. Start the timer
timer clear 1
timer on 1

// 3. Start the simulation loop
forvalues i = 1/500 {

    quietly {
        clear
        set obs 525000

        gen famid = _n
        gen u = rnormal()

        // Use expand 2 instead of reshape long
        expand 2
        bysort famid: gen sib_rank = _n

        // Generate exposure and outcome directly
        gen exposure = rbinomial(1, invlogit(logit(.48) + log(4)*u)) if sib_rank==1
        replace exposure = rbinomial(1, invlogit(logit(.50) + log(4)*u)) if sib_rank==2

        gen outcome = rbinomial(1, invlogit(logit(.0125) + log(1)*exposure + log(4)*u)) if sib_rank==1
        replace outcome = rbinomial(1, invlogit(logit(.0075) + log(1)*exposure + log(4)*u)) if sib_rank==2

        // Keep only discordant exposure pairs
        bysort famid: egen exp_sum = sum(exposure)
        keep if exp_sum == 1
```

```

        // Count retained pairs
        local n_pairs = _N / 2

        // Define sets for the conditional models
        bysort famid (sib_rank): gen is_set_A = (exposure[1] == 1)

        // Estimate models and store the exponentiated odds ratios
        clogit outcome exposure if is_set_A==1, or group(famid)
        local or_1 = exp(_b[exposure])

        clogit outcome exposure if is_set_A==0, or group(famid)
        local or_2 = exp(_b[exposure])

        clogit outcome exposure , or group(famid)
        local or_3 = exp(_b[exposure])

        clogit outcome exposure sib_rank, or group(famid)
        local or_4 = exp(_b[exposure])

        // Post the extracted values to the file
        post mypost (`or_1') (`or_2') (`or_3') (`or_4') (`n_pairs')
    }

    // Print progress to the console
    if mod(`i', 50) == 0 {
        noisily display "Completed iteration `i'"
    }
}

// 4. Stop the timer
timer off 1

// 5. Close the postfile
postclose mypost

// 6. Load the aggregated results and display the final summaries
use `sim_results', clear

display ""
display "--- Mean Number of Discordant Exposure Pairs Retained ---"
summarize n_discordant, meanonly
display "Mean retained pairs: " round(r(mean), 1)
display ""

display "--- Median Odds Ratios and Empirical 95% CIs (500 runs) ---"
display "---------------------------------------------------------"
display "Variable       |      2.5% |       50% |     97.5%"
```

```
display "---------------------------------------------------------"
foreach var in or_first or_second or_all or_adjustrank {
    quietly centile `var', centile(2.5 50 97.5)
    display %-14s "`var'" " | " %9.6f r(c_1) " | " %9.6f r(c_2) " | " %9.6f r(c_3)
}
display "---------------------------------------------------------"
display ""

// 7. Display the total execution time
quietly timer list 1
local minutes = floor(r(t1) / 60)
local seconds = round(mod(r(t1), 60), 0.1)
display "--- Simulation Execution Time ---"
display "Total time: `minutes' minutes and `seconds' seconds"


```
